# Tumor Biology and Patterns of Recurrence in High-Grade Glioma: Implications for Radiation Target Delineation

**DOI:** 10.64898/2026.04.23.26351633

**Authors:** Rahul Barve, Darshan Gowda, K. Jeyaram Illiayaraja

**Affiliations:** Department of Radiation Oncology, British Columbia Cancer Agency, Abbotsford, BC, Canada; Department of Radiation Oncology, Max Super Specialty Hospital, Saket, New Delhi, India; Department of Medicine, Hassan Institute of Medical Sciences, Hassan, India; Biostatistics, Strategy Planning and Health Intelligence (SPHI), Primary Health Care Corporation (PHCC), Doha, Qatar

**Keywords:** Radiotherapy, High-grade glioma, Glioblastoma, Pattern of recurrence, Radioresistance, Tumor biologys

## Abstract

**Purpose:** Recurrence in high-grade glioma (HGG) predominantly occurs within the high-dose radiation field, raising the question of whether treatment failure reflects limitations in radiation target delineation or is driven by intrinsic tumor biology. This study evaluated recurrence patterns following standard chemoradiotherapy and their treatment implications.

**Material and Methods:** This retrospective single-center study included 41 patients with histologically confirmed HGG treated with surgery followed by radiotherapy with concurrent and adjuvant temozolomide (TMZ). Patients were followed through August 2018; those with recurrence were included in the analysis. Recurrence patterns were classified based on their spatial relationship to the 60 Gy isodose line as central, in-field, marginal, or distant. Survival outcomes were estimated using the Kaplan–Meier method and compared using the log-rank test.

**Results:** The most common pattern of recurrence was central (15 patients, 36.5%), followed by in-field (11, 26.8%), distant (6, 14.6%), marginal (5, 12.1%), and multicentric (4, 9.8%). Central and in-field recurrences (local failures) accounted for 26 patients (63%). Median overall survival (OS) was 27 months, and median progression-free survival (PFS) was 12 months. Survival differed significantly by recurrence pattern (log-rank p = 0.018), with marginal recurrence associated with more favorable outcomes.

**Conclusion:** The predominance of central and in-field recurrences within the high-dose region suggests that treatment failure in HGG is not solely explained by inadequate target delineation and may also be driven, in part, by intrinsic tumor biology, including radioresistant subpopulations and tumor heterogeneity. Future strategies may benefit from incorporating biologically guided approaches alongside optimization of radiation treatment parameters.

## Introduction

High-grade gliomas, particularly glioblastoma, are among the most aggressive primary brain tumors and are characterized by rapid proliferation, diffuse infiltration, and poor prognosis (1). In this study, the term high-grade glioma (HGG) refers to aggressive diffuse gliomas treated during the study period, acknowledging that the WHO 2021 classification distinguishes IDH-wildtype glioblastoma from IDH-mutant astrocytoma, grade 4 (2). Despite advances in neurosurgical techniques and imaging, complete resection remains challenging because of the infiltrative nature of these tumors. The current standard of care consists of maximal safe resection followed by radiotherapy with concurrent and adjuvant temozolomide (TMZ), with the addition of tumor-treating fields in selected patients (3). However, survival outcomes have improved only modestly, with median overall survival of approximately 14–16 months with standard chemoradiation and up to approximately 20 months in studies incorporating tumor-treating fields (4,5).

A defining feature of HGG is its pattern of recurrence. Multiple studies have shown that most recurrences arise within the high-dose radiation field, typically in central or in-field locations (6,7). This pattern persists despite advances in surgical techniques, imaging, and radiation planning, raising important questions about the mechanisms underlying treatment failure.

Historically, recurrence patterns have been interpreted primarily in relation to radiation target delineation, including the adequacy of clinical target volume margins and the possibility of geographic miss. However, the continued predominance of in-field recurrences despite appropriate target coverage suggests that factors beyond treatment delivery may contribute significantly.

Increasing evidence points toward a role for intrinsic tumor biology in driving treatment resistance. Glioma stem-like cells, tumor hypoxia, and microenvironmental influences have been implicated in promoting radioresistance and tumor repopulation following therapy (8,9). These mechanisms may help explain the limited benefit observed with dose-escalation strategies and the persistence of viable tumor cells within irradiated regions (10,11).

In this context, the present study aims to evaluate patterns of recurrence following standard chemoradiotherapy in patients with HGG and to examine whether these patterns are more consistent with limitations in radiation target delineation or with underlying tumor biology, with the goal of informing future therapeutic approaches.

## Materials and Methods

### Study Design and Patient Population

This retrospective, single-center, non-randomized study was conducted at Max Cancer Centre, Saket, New Delhi, India. Patients with high-grade glioma (HGG) who underwent gross total tumor resection followed by standard radiotherapy with concurrent temozolomide (TMZ) and subsequent adjuvant TMZ were included. Patients who received primary treatment between January 2012 and December 2016 and subsequently developed recurrence during follow-up (through April 2018) were eligible for pattern-of-failure analysis.

A total of 41 patients were retrospectively identified and included in the analysis. Eligible patients were aged 18–70 years. Functional status was assessed using the Karnofsky Performance Status (KPS), a standardized scale ranging from 0 to 100 that evaluates a patient’s ability to perform daily activities, with higher scores indicating better functional status (12). Inclusion criteria comprised KPS ≥50, histologically confirmed high-grade glioma (WHO grade IV, glioblastoma), and prior gross total tumor resection. Patients were excluded if they had received prior radiotherapy outside the institution, had low-grade gliomas, underwent biopsy only or subtotal resection, or were outside the specified age range.

Molecular markers, including IDH mutation status, were not routinely available during the study period and were therefore not included in the analysis.

### Ethical Considerations

This study was approved by the Institutional Review Board (Approval No: IEC/2025/ONC/112). The requirement for informed consent was waived due to the retrospective nature of the study and use of de-identified data.

### Treatment Details

#### Radiotherapy

As this was a retrospective study, all patients had previously received standard-of-care treatment, and no interventions were performed as part of the study. Patients were followed longitudinally with serial imaging to evaluate patterns of recurrence. Details of the radiotherapy technique are described below.

Radiotherapy was initiated within 4 weeks following surgery. Planning computed tomography (CT) images were co-registered with pre- and postoperative contrast-enhanced magnetic resonance imaging (MRI), including T1-weighted, T2-weighted, and FLAIR sequences, for target delineation.

Target volumes were defined as follows: the gross tumor volume (GTV) encompassed the contrast-enhancing tumor on T1-weighted MRI, while the clinical target volume (CTV) included the GTV with surrounding edema identified on T2/FLAIR sequences. Planning target volumes (PTVs) were generated by applying appropriate margins. PTV2 was defined as the CTV with a 1.5 cm margin and treated to 54–55 Gy, followed by a sequential boost to PTV1, defined as the GTV with a 0.5 cm margin, to a total dose of 60 Gy in 30 fractions (Image 1).

All patients received external beam radiotherapy using 6 MV photon beams delivered via a linear accelerator with intensity-modulated radiation therapy (IMRT).

### Chemotherapy

Concurrent chemotherapy consisted of temozolomide (TMZ) administered at 75 mg/m^2^ daily throughout radiotherapy. Adjuvant TMZ was initiated 4 weeks after completion of radiotherapy and administered for 5 consecutive days every 28 days. The initial dose was 150 mg/m^2^, escalated to 200 mg/m^2^ from cycle 2 if tolerated. Adjuvant TMZ was continued for up to 12 cycles or until disease progression or unacceptable toxicity.

### Data Collection

Clinical and treatment data were obtained from outpatient records, radiotherapy planning systems, and dose–volume histogram (DVH) databases. All data were anonymized and recorded in a structured data collection form.

### Imaging and Assessment of Recurrence

Patients were followed with clinical evaluation and contrast-enhanced magnetic resonance imaging (MRI) approximately 4 weeks after completion of radiotherapy, every 2–3 months during the first 2 years, and at increasing intervals of 3–6 months thereafter, or earlier if clinically indicated.

Tumor recurrence was defined according to the Response Assessment in Neuro-Oncology (RANO) criteria as any of the following: a ≥25% increase in the sum of the products of perpendicular diameters of enhancing lesions, the appearance of a new lesion, or clinical deterioration (13). Pseudoprogression was defined as transient radiological worsening occurring within 2–6 months after completion of chemoradiation that subsequently stabilized or resolved on follow-up imaging.

All imaging studies were independently reviewed by two experienced neuroradiologists. In cases of diagnostic uncertainty, follow-up imaging and clinical correlation were used to distinguish true progression from treatment-related changes. Discrepancies between reviewers were resolved by consensus through joint image review, incorporating clinical findings and serial imaging to confirm true progression.

### Recurrence Pattern Analysis

Recurrent tumor volumes (Vrecur) were contoured on fused CT MRI datasets. Recurrent tumor volumes were contoured by the attending radiation oncologist, with neuroradiology input provided through direct consultation or multidisciplinary tumor board image review, and final contouring decisions were reached by consensus Recurrence patterns were classified based on spatial relation to the 60 Gy isodose line (IDL): Central recurrence, >95% of the Vrecur inside the 60-Gy IDL In-field recurrence, 80-95% of the Vrecur inside the 60-Gy IDL Marginal recurrence, 20-80% of the Vrecur inside the 60-Gy IDL Distant recurrence, <20% of the Vrecur inside the 60-Gy IDL Multicentric recurrence: presence of two or more non-contiguous lesions separated by normal brain tissue, irrespective of their relation to the 60 Gy IDL

### Statistical Analysis

Overall survival (OS), progression-free survival (PFS), and time to recurrence were estimated using the Kaplan–Meier method. Overall survival was defined as the time from diagnosis to death from any cause or last follow-up. Progression-free survival was defined as the time from diagnosis to radiological progression or death. Time to recurrence was defined as the interval from completion of radiotherapy to radiologically confirmed recurrence. Patients who were alive or progression-free at last follow-up were censored. Survival differences between groups were compared using the log-rank test. A two-sided p-value <0.05 was considered statistically significant. Statistical analyses were performed using STATA version 17 (StataCorp LLC, College Station, TX, USA).

## Results

### Patient Cohort and Follow-up

A total of 41 patients with high-grade glioma were included in the analysis. Baseline characteristics of the study cohort are summarized in Table 1. At the time of analysis, 6 patients were alive and 35 had died. The median follow-up duration was 24 months. The median time to radiological recurrence from completion of radiotherapy was 12 months.

**Table 1.** Baseline clinicopathologic characteristics of patients with recurrent (n = 41)

| Characteristics | Patients |
| --- | --- |
| **Gender** |  |
| Male | 30 |
| Female | 11 |
| **Pathological diagnosis** |  |
| High Grade Glioma<br>(GBM or Astrocytoma grade 4) | 41 |
| **Age (years)** |  |
| Median | 52 |
| Range | 27-70 |
| **Karnofsky performance status** |  |
| Median | 80 |
| Range | 50- 90 |

### Survival Outcomes

For the entire cohort, the median overall survival (OS) was 27 months. The 1-year and 2-year OS rates were 85.3% and 63.4%, respectively. The median progression-free survival (PFS) was 12 months.

### Patterns of Recurrence

All patients developed intracranial recurrence, as confirmed on serial MRI. The spatial patterns of recurrence, determined by co-registration of recurrence imaging with the original radiotherapy plan, are illustrated in Figures 2–5.

**Figure 1.**
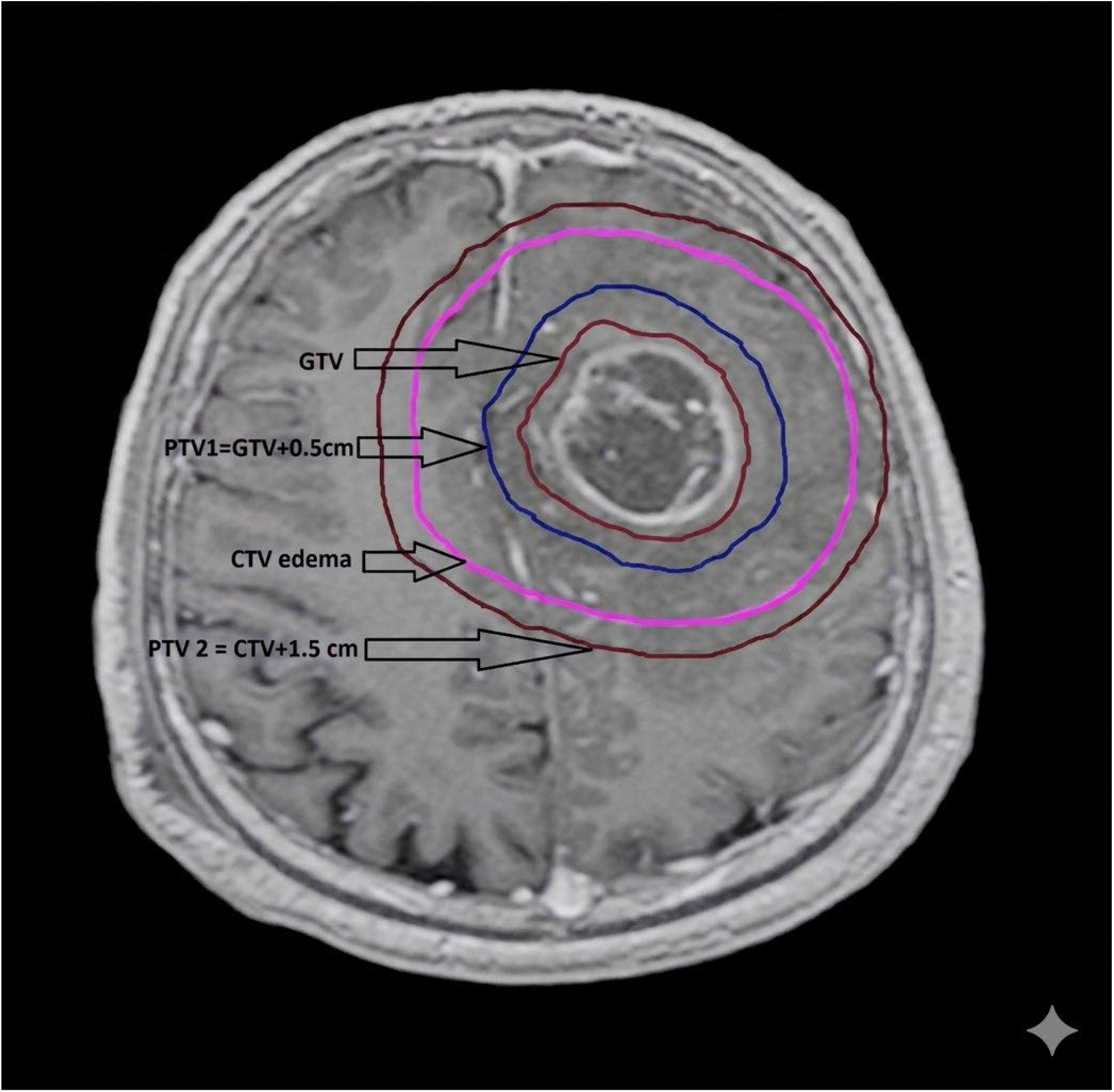
Radiotherapy target volume delineation for HGG. Representative axial post-contrast T1-weighted MRI demonstrating GTV, PTV1 (GTV + 0.5 cm), and PTV2 (CTV + 1.5 cm including peritumoral edema).

**Figure 2.**
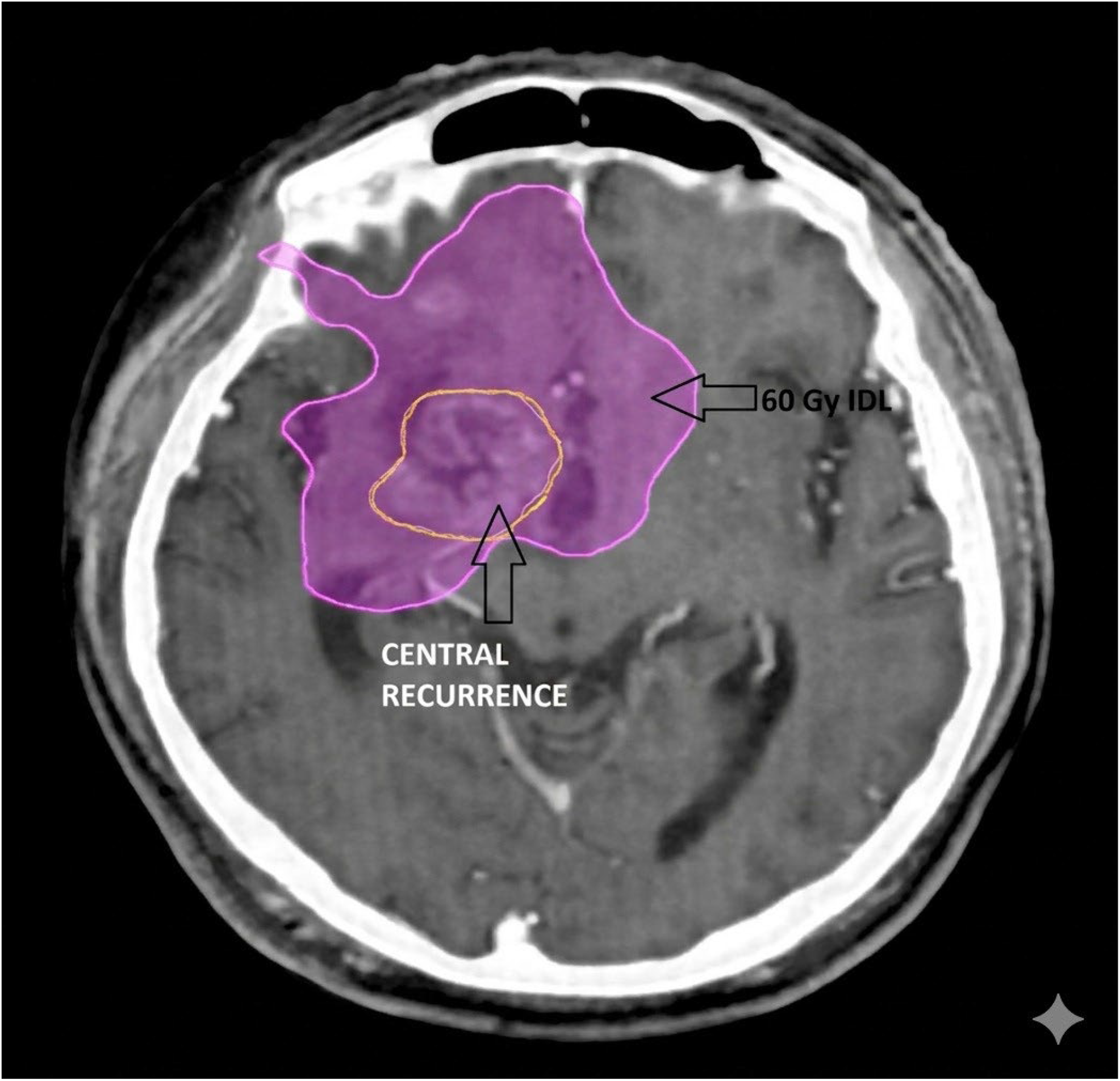
Central recurrence. Representative example of central recurrence, with >95% of the recurrent lesion (Vrecur) located within the 60-Gy isodose line (IDL)

The most common pattern of first recurrence was central recurrence, observed in 15 patients (36.5%) (Figure 2). This was followed by in-field recurrence in 11 patients (26.8%) (Figure 3), marginal recurrence in 5 patients (12.1%) (Figure 4), distant recurrence in 6 patients (14.6%) (Figure 5), and multicentric recurrence in 4 patients (9.8%). Central and in-field recurrences were classified as local failures and together accounted for 26 patients (63%) of all recurrences. The patterns of recurrence and associated survival outcomes are summarized in Table 2.

**Table 2.**
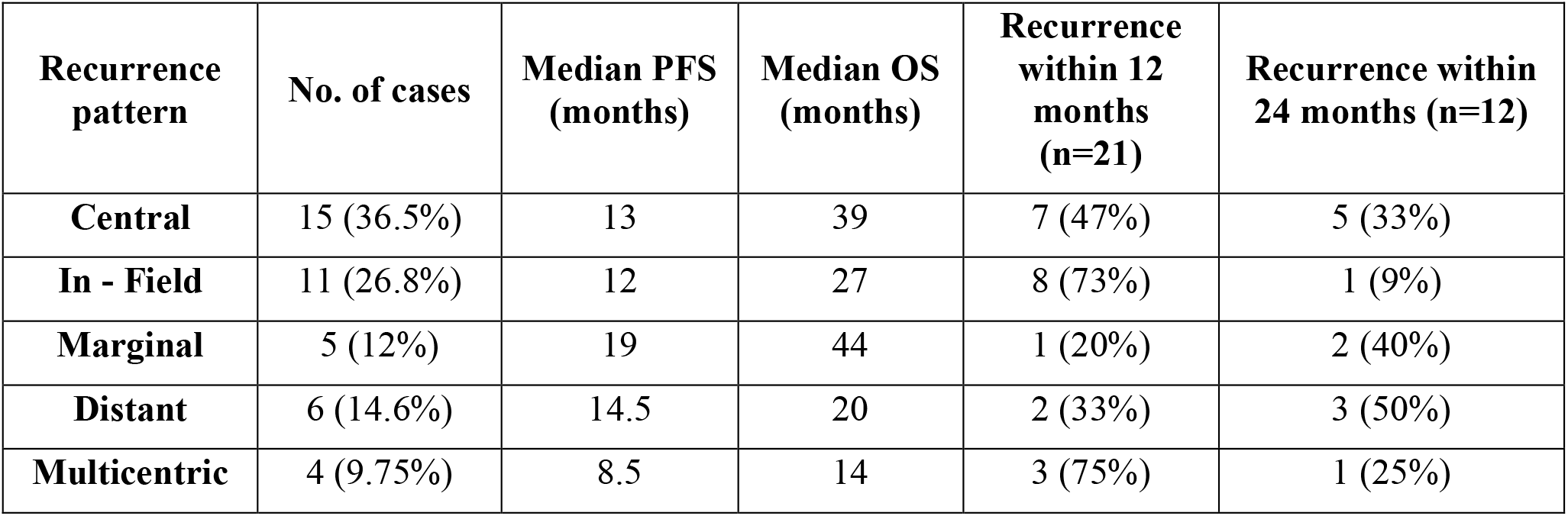
Recurrence pattern and survival outcomes (n = 41)

**Figure 3.**
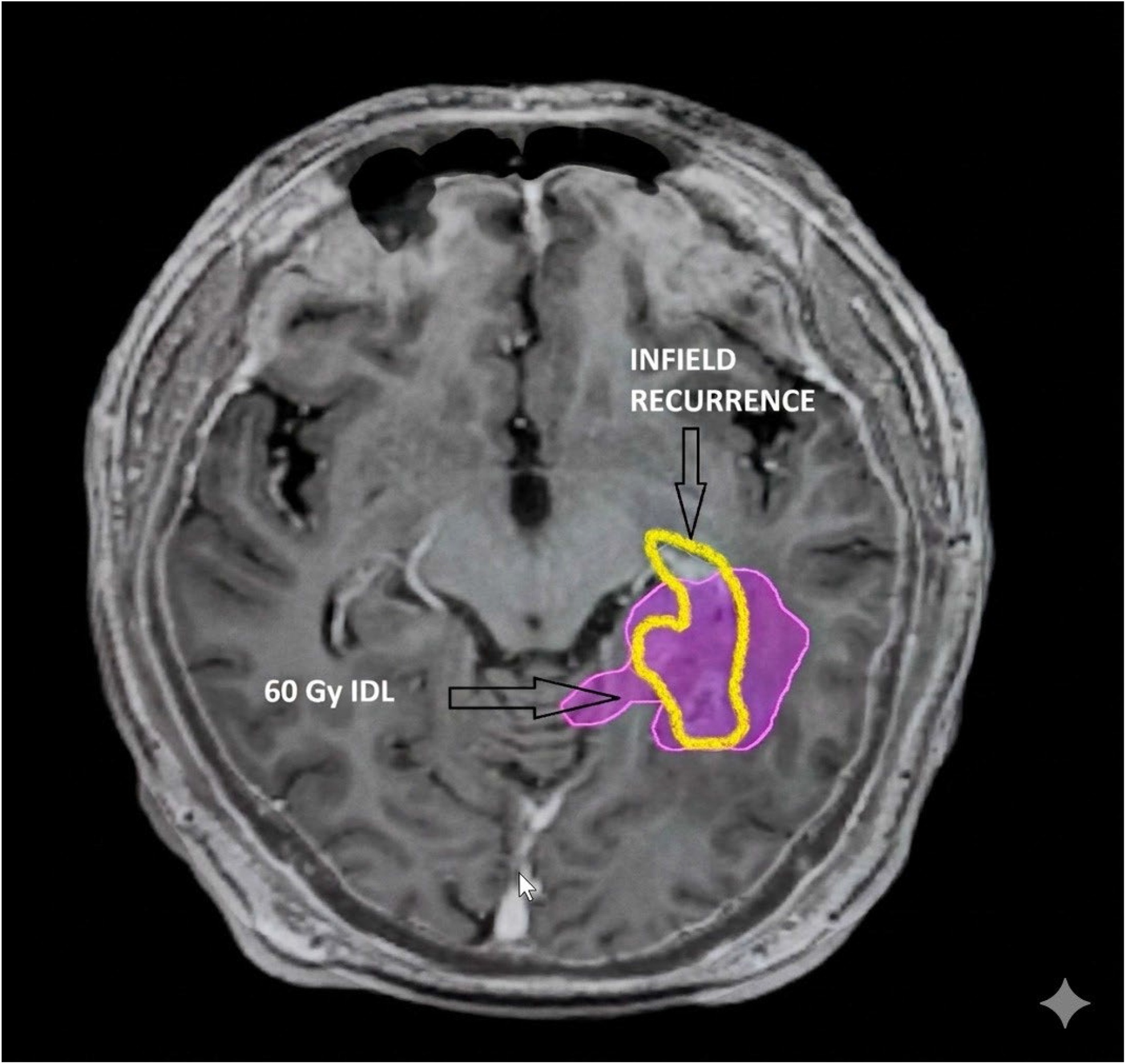
In-field recurrence. Representative example of infield recurrence, with 80-95% of the recurrent lesion (Vrecur) located within the 60-Gy isodose line (IDL)

**Figure 4.**
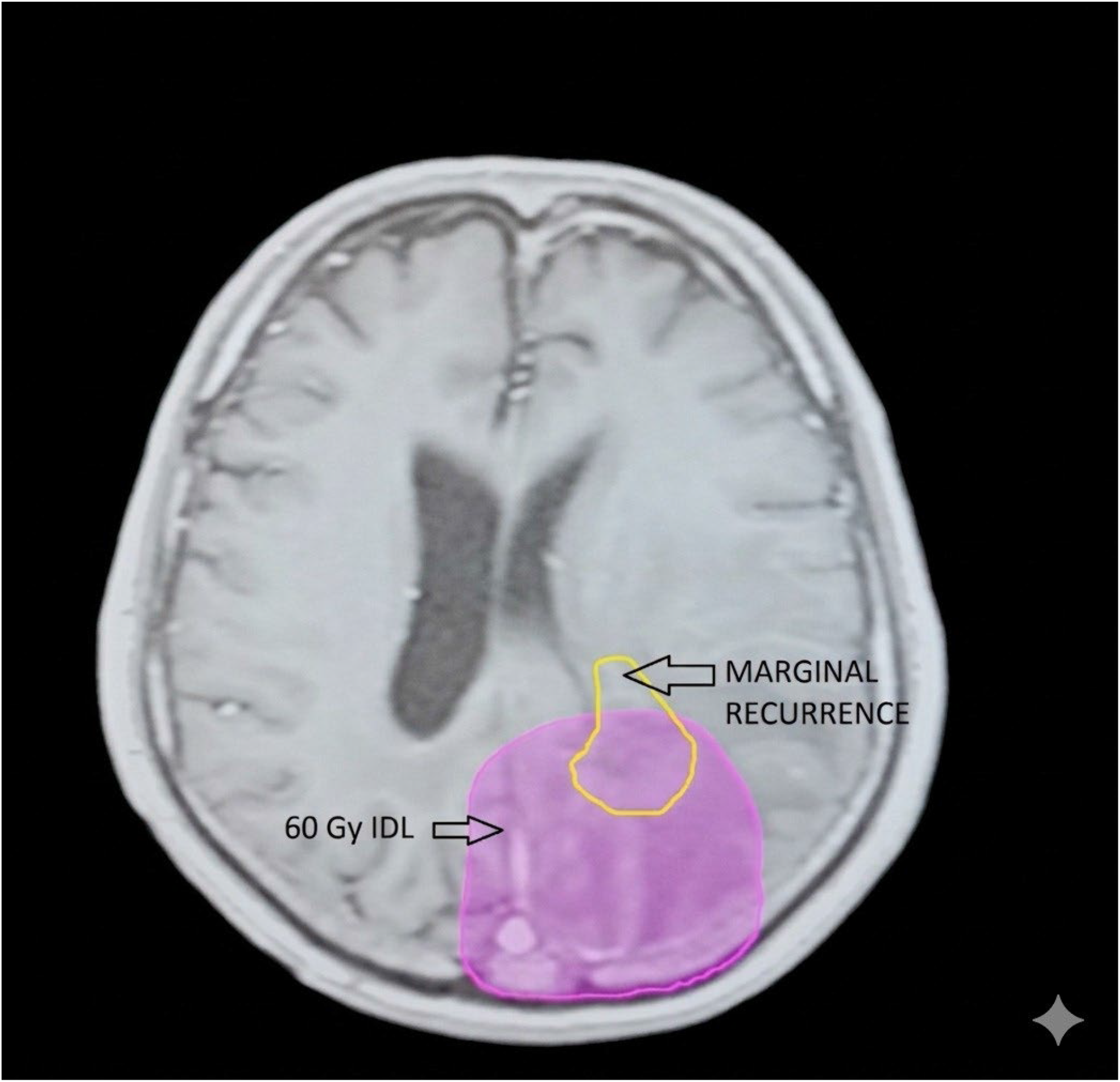
Marginal recurrence. Representative example of marginal recurrence, with 20-80% of the recurrent lesion (Vrecur) located within the 60-Gy isodose line (IDL)

**Figure 5.**
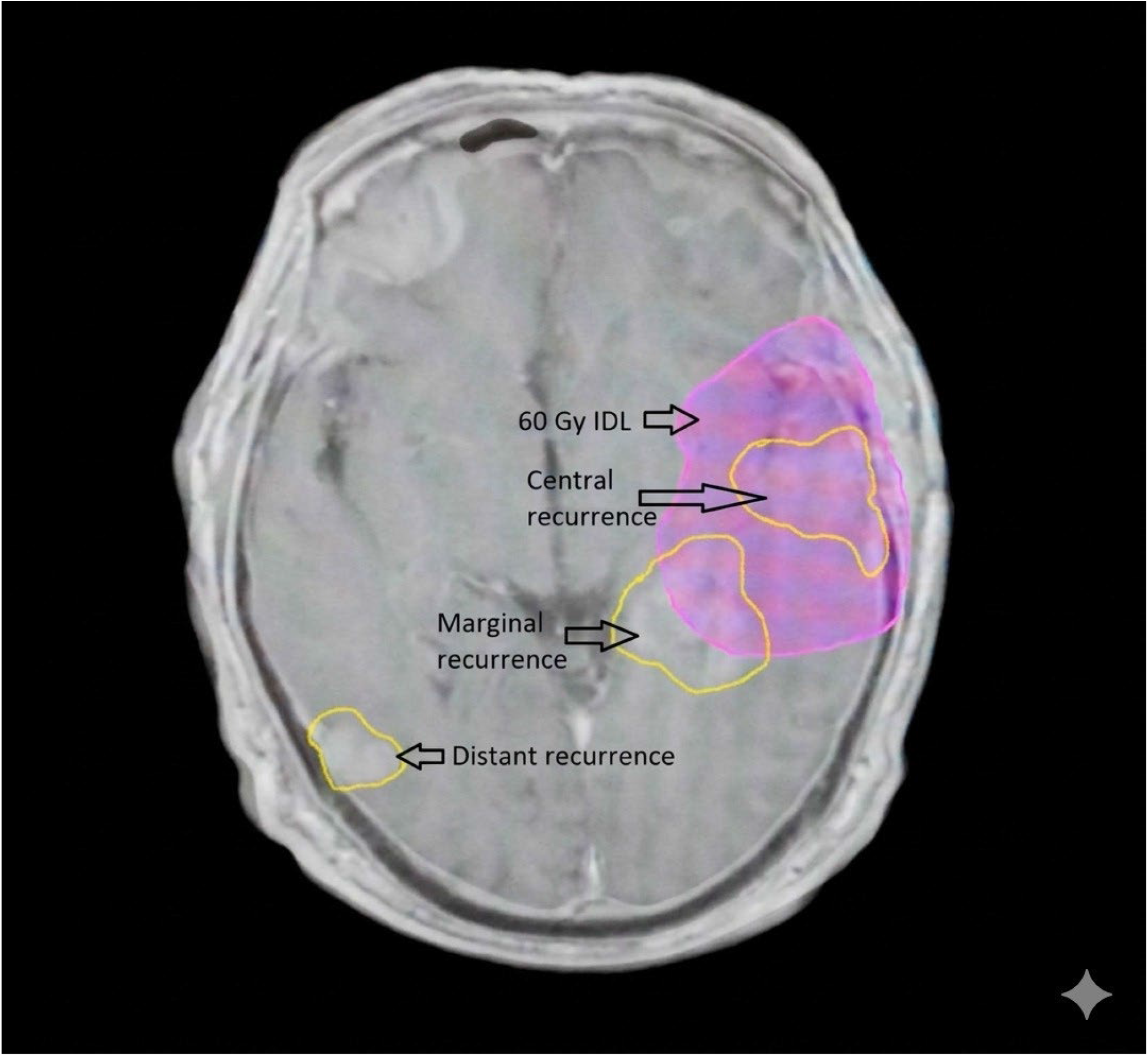
Illustrative examples of recurrence patterns relative to the 60 Gy IDL. Representative axial MRI showing examples of central, marginal, and distant recurrence in relation to the 60 Gy isodose line.

Patients with central recurrence demonstrated a median OS of 39 months, while those with in-field recurrence had a median OS of 27 months. Marginal recurrence was associated with the most favorable outcomes, with a median OS of 44 months. In contrast, distant and multicentric recurrences were associated with poorer outcomes, with median OS of 20 months and 14 months, respectively.

Kaplan–Meier survival analysis demonstrated a statistically significant difference in overall survival according to recurrence pattern (log-rank p = 0.018) (Figure 6).

**Figure 6.**
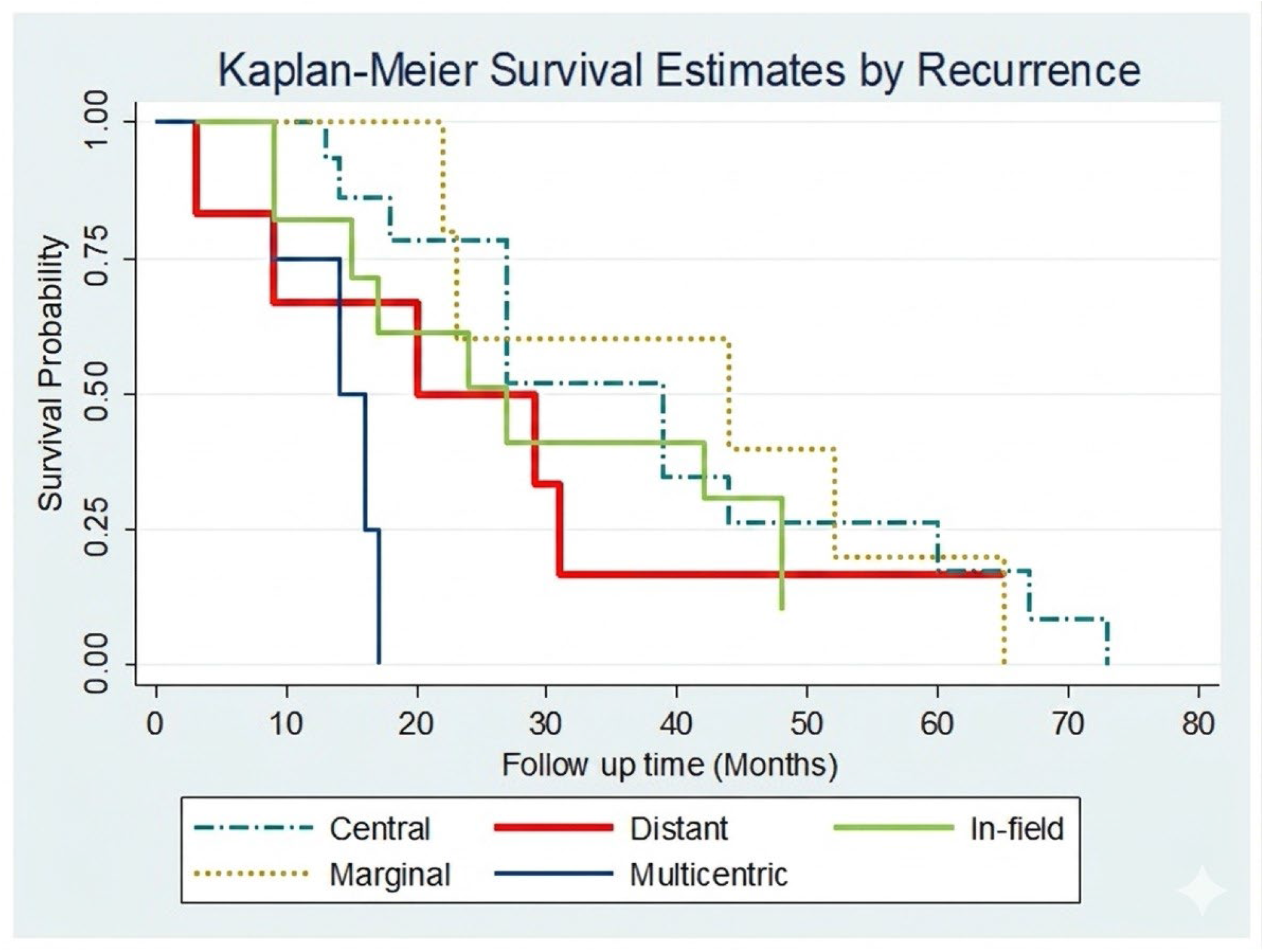
Kaplan-Meier analysis of overall survival according to recurrence pattern. Kaplan-Meier curves comparing overall survival across recurrence patterns. Survival distributions differed significantly by recurrence pattern (log-rank p = 0.018).

### MGMT Promoter Methylation Status

MGMT promoter methylation status was available for 23 patients (56%). Among these, 14 patients were methylated and 9 were unmethylated. Patients with MGMT-methylated tumors demonstrated longer median overall survival (39 months vs. 27 months); however, given the limited sample size and incomplete data availability, this finding should be interpreted with caution.

### Karnofsky Performance Status (KPS)

Karnofsky Performance status was a significant predictor of survival. Patients with KPS >70 had a median OS of 31 months, compared with 20 months in those with KPS ≤70 (log-rank p = 0.01)

## Discussion

In the present study, the proportion of central and in-field recurrences (63%) was lower than that reported in historical series (approximately 80%), which may reflect the small sample size and cohort heterogeneity. Nevertheless, the majority of recurrences occurred within the high-dose radiation region, consistent with prior reports (6,7). This finding suggests that treatment failure in high-grade glioma is not solely attributable to limitations in radiation target delineation and may reflect underlying tumor biology.

In a retrospective analysis of 207 patients with glioblastoma, Minniti et al. (14) reported predominantly in-field recurrences (87%), with only 2.4% marginal failures using a 2 cm clinical target volume (CTV) margin. Notably, simulated reduction to a 1 cm margin did not alter recurrence patterns while significantly reducing normal brain dose, supporting the feasibility of margin de-escalation. Similarly, Braschi et al. (15) reported outcomes using a 1 cm CTV margin, with only 2% marginal recurrence, further suggesting that larger margins may not substantially reduce the risk of treatment failure.

Dose escalation strategies in glioblastoma have not consistently improved outcomes. The phase III SPECTRO GLIO trial (Laprie et al., 2024) (10) demonstrated no improvement in overall or progression-free survival with magnetic resonance spectroscopic imaging (MRSI)-guided dose escalation to 72 Gy compared with standard 60 Gy, without increased toxicity. Similarly, a meta-analysis of 22 prospective trials involving over 2,000 patients showed that while dose-escalated radiotherapy alone improved outcomes compared with radiotherapy alone, this benefit was not observed when combined with temozolomide, irrespective of MGMT status (11). Although several phase II studies using advanced imaging modalities, including MR spectroscopy, perfusion MRI, and amino acid PET, have reported encouraging results with focal dose escalation, these findings remain unconfirmed in randomized trials (16,17).

Taken together, these findings highlight an important clinical gap, namely whether recurrence patterns observed in earlier cohorts continue to justify large treatment margins in the modern era of molecularly informed care. Current evidence suggests that dose escalation alone is unlikely to overcome intrinsic radioresistance in glioblastoma. In this context, our findings support the need to consider biologically informed strategies alongside continued refinement of radiation target delineation.

Emerging evidence underscores the biological complexity of glioblastoma. Intratumoral heterogeneity results in distinct cellular subclones with variable sensitivity to radiation and chemotherapy (18). In particular, glioma stem-like cells, which reside within hypoxic and perivascular niches, demonstrate enhanced DNA damage repair and resistance to apoptosis, enabling tumor repopulation following treatment (9,19). In addition, the tumor microenvironment, characterized by hypoxia, angiogenesis, and immune suppression, further contributes to treatment resistance by limiting the effectiveness of radiation-induced DNA damage and promoting adaptive survival pathways (20). These mechanisms provide a biological framework that may explain the persistence of predominantly in-field recurrences despite adequate radiation delivery and the limited benefit of dose escalation or margin expansion (10,21). Collectively, these findings support a biologically driven pattern of recurrence (Table 3).

**Table 3.** Biological mechanisms underlying local recurrence in HGG. Source: Table created by the authors based on data extracted and synthesized from the cited studies.

| Biological Finding | Summary | Key Evidence |
| --- | --- | --- |
| Residual infiltrative tumor cells at the resection margin | Tumor cells extend beyond the contrast-enhancing margin on MRI and contribute to central or in-field recurrence. | Spiteri I et al., 2019; Li X et al., 2025 (22,23) |
| Glioma stem-like cells (GSCs) | GSCs exhibit enhanced DNA repair capacity, resistance to apoptosis, and persistence within protective niches, enabling tumor repopulation after treatment. | Bao S et al., 2006; Al-Holou WN et al., 2023 (19,24) |
| Radiation-induced microenvironmental changes | Radiation may promote a pro-tumorigenic microenvironment, enhancing tumor survival, invasion, and mesenchymal transition | Fletcher-Sananikone E et al., 2021; Yoo KC et al., 2018 (25,26) |
| VC-Resist (Vessel co-opting mediated resistant) | Following treatment, GBM cells may migrate to perivascular niches, where survival signaling promotes resistance and local recurrence. | Pichol-Thievend C et al., 2024 (27) |
| Invasive cell resistance | Peripheral invasive tumor cells demonstrate enhanced DNA repair, reduced apoptosis, and transcriptional adaptations associated with treatment resistance. | Tang Z et al., 2022 (28) |
| Clonal evolution under therapy | Recurrent GBM may arise from pre-existing local subclones (e.g., TP53-mutant GSCs) that are positively selected under therapeutic pressure rather than representing de novo distant origin. | Orzan F et al., 2017 (29) |

Beyond local therapy, the limited efficacy of systemic treatments further highlights the role of tumor biology in glioblastoma. Immune checkpoint inhibitors, including anti–PD-1 therapies, have not demonstrated a survival benefit in either newly diagnosed or recurrent disease, likely reflecting an immunologically “cold” tumor microenvironment, low tumor mutational burden, and significant immunosuppression (30,31). Similarly, targeted therapies such as CDK4/6 inhibitors, despite a strong biological rationale, have shown limited clinical efficacy as monotherapy, with most patients experiencing early disease progression (32–34).

Emerging therapeutic approaches, including PARP inhibitors (35,36), dendritic cell vaccines (37), and adoptive cellular therapies such as CAR-T cells (38,39), have demonstrated encouraging signals in early-phase studies but remain investigational. These strategies may offer benefit in selected patient populations, particularly when integrated with standard treatment modalities.

Taken together, our findings suggest the need to reconsider current treatment strategies in high-grade glioma. Rather than focusing solely on expansion of radiation margins or dose escalation, future approaches may benefit from incorporating biologically guided strategies, including molecular profiling, targeted systemic therapies, and immunomodulatory approaches, alongside continued refinement of radiation treatment parameters such as dose and target delineation, to better address the underlying drivers of treatment resistance.

### Limitations

This study is limited by its retrospective design and small, single-institution cohort, which may limit generalizability and preclude definitive biological correlations. In addition, the study reflects an earlier treatment era and does not incorporate contemporary molecular classification. The analysis is based on imaging-defined recurrence patterns, which may be subject to registration uncertainty and interobserver variability. However, the findings remain clinically relevant, as standard chemoradiation continues to be the backbone of treatment and local recurrence remains the predominant pattern of failure, supporting a role for underlying tumor biology in treatment resistance.

### Future Directions

Future research should focus on addressing the biological drivers of treatment resistance in high-grade glioma, particularly glioblastoma. Novel therapies, including PARP inhibitors, CDK4/6 inhibitors, dendritic cell vaccines, and CAR-T cell therapies, have shown promising early signals but remain investigational. Approaches such as MRI-guided reirradiation highlight the potential of adaptive, image-guided dose delivery, although their impact on survival outcomes remains to be established (40). Integration of molecular profiling with biologically guided radiotherapy may enable patient-specific treatment adaptation and improved risk stratification.

## Conclusion

In this retrospective cohort, recurrence in HGG occurred predominantly within the high-dose radiation region, with central and in-field failures accounting for the majority of cases. These findings highlight the persistent challenge of local disease control and suggest that treatment failure is not solely attributable to target delineation but is also likely influenced by intrinsic radioresistant tumor biology. This supports the need for treatment strategies that extend beyond conventional chemoradiation, including biologically informed and targeted approaches. Despite current advances, participation in clinical trials remains an important consideration for eligible patients with both newly diagnosed and recurrent HGG.Author

## Contributions

All authors have reviewed the final version to be published and agreed to be accountable for all aspects of the work.

Concept and design: Rahul Barve

Acquisition, analysis, or interpretation of data: Rahul Barve, Darshan S. Gowda, K.Jeyaram Illiayaraja

Drafting of the manuscript: Rahul Barve, Darshan S. Gowda

Critical review of the manuscript for important intellectual content: Rahul Barve, Darshan S. Gowda, K.Jeyaram Illiayaraja

## Data Availability Statement

The datasets generated and/or analyzed during the current study are not publicly available due to institutional policies but are available from the corresponding author on reasonable request.

